# Translational impact of basic science promotes a new era of precision medicine for migraine

**DOI:** 10.1101/2023.07.25.23293169

**Authors:** Frank Porreca, Edita Navratilova, Joe Hirman, Antoinette MaassenVanDenBrink, Richard B. Lipton, David W. Dodick

**Author notes:** **Correspondence:** Frank Porreca, Ph.D., Department of Pharmacology, University of Arizona, Tucson, AZ 85724 USA.

## Abstract

Precision medicine has emerged as a powerful approach to improve treatment outcomes for many medical conditions by consideration of the genetic characteristics of the patient. Migraine is likely to be the world’s most common neurological disorder affecting over 1 billion people, approximately 700 million of whom are women. Yet, patient sex, the most basic genetic difference, is rarely considered in selection of therapies for people with migraine. Preclinical studies reveal that calcitonin gene-related peptide (CGRP), a neurotransmitter thought to be causal in promoting migraine in many patients, elicits female selective pain and headache-like responses. Consistent with this, we report a subgroup analysis of publicly available clinical data evaluating small molecule CGRP receptor antagonists for the acute migraine revealing preferential efficacy in women. Analyses of human post-mortem sensory neurons from male and female donors reveal sexual dimorphism at transcript, protein and functional levels. These findings raise the possibility that regardless of common diagnosis and phenotype, mechanisms promoting migraine pain may differ between sexes. Such qualitative sex differences suggest that clinical trials should include sex-specific analyses, that sex stratified treatment guidelines may improve treatment outcomes in migraine and that the uniform therapeutic approach to pain-related disorders in men and women requires reassessment.

---

Maintaining the highest standard of patient care requires constant updating of established practice as new information on disease pathophysiology emerges. Migraine is a multi-symptom, multiphasic and chronic neurological disorder that impairs the quality of life of patients around the world. Options for treatment of migraine now include drugs that interfere with calcitonin gene-related peptide (CGRP) signaling.^1^ The gepant class of molecules are CGRP receptor (CGRP-R) antagonists and include ubrogepant, rimegepant and zavegepant used for acute migraine therapy. Multiple lines of evidence, including the clinical effectiveness of the gepant drugs, suggest a causal role of CGRP in migraine pathophysiology. However, CGRP-R antagonists are not effective in all patients for acute migraine treatment. Understanding which patient groups would preferentially respond to CGRP-R antagonists would improve patient care and avoid the use of these drugs in likely non-responders.

For reasons that are not understood, most migraine patients are women.^2^ Female sex hormones have been recognized as factors in promoting migraine. While CGRP likely promotes migraine in some patients, whether this mechanism applies equally across sexes is unknown. Women show increased severity, persistence and co-morbidity profiles as well as increased prevalence of migraine relative to men.^3,4^ These observations raise the possibilities that (a) CGRP may preferentially promote migraine in women and (b) treatment effects of CGRP-R antagonists may not be uniform in men and women. Review of data from clinical trials of gepant drugs, as well as data from preclinical studies and human post-mortem tissues, may provide insight into these possibilities.

## Evidence for CGRP in promoting migraine

Three crucial lines of evidence support the contribution of CGRP as a migraine substrate in humans: (a) increased levels of this transmitter were observed in the jugular outflow of people with migraine during attacks and normalization of CGRP levels was observed following relief of headache pain with sumatriptan;^5^ (b) infusion of CGRP in people with migraine provokes headache with a migraine phenotype^6^ and (c) therapies that prevent CGRP signaling are clinically effective. Critically, however, evidence supporting CGRP was obtained from studies conducted almost exclusively in women limiting conclusions regarding the causal role of CGRP across sexes.

The initial study demonstrating elevated CGRP levels during migraine and normalization by sumatriptan was conducted in 8 patients, 7 of whom were women.^5^ While all of the female patients showed elevated CGRP levels during migraine, responded to sumatriptan, and returned to baseline CGRP levels after treatment, the one male patient did not exhibit elevated CGRP during migraine, did not respond to sumatriptan and CGRP levels remained unchanged after treatment.^5^ Thus, the conclusions of this seminal study should be limited to women. Provocative studies of CGRP infusion to elicit migraine have also been conducted almost exclusively in women.^7^ Such studies must be interpreted with caution as the pharmacological dose of CGRP used to elicit migraine may not reflect the contribution of endogenous CGRP in the trigeminal system during naturally occurring migraine. The clinical effectiveness of the gepants for acute migraine therapy provides the most convincing evidence for a causal role of CGRP in this disorder. To date, however, detailed analysis of the efficacy of CGRP-R antagonists for treatment of acute migraine by sex has not been published.

### Effectiveness of CGRP-R antagonists for acute treatment of migraine in men and women

Clinical and statistical reviews by the FDA Center for Drug Evaluation and Research (CDER) for the New Drug Applications (NDA) of ubrogepant,^8^ rimegepant,^9^ and zavegepant^10^ in the acute treatment of migraine are publicly available. We conducted a subpopulation analysis of available published data from the CDER reviews to evaluate potential sex differences in the response rates of these three gepants. Results from each confirmatory study used to support a labeled dose level were used. Drug response (% treatment effect) was determined by subtracting the response to placebo from the response to treatment on two co-primary endpoints (a) the 2-hour pain freedom (PF) (Table 1) and (b) the 2-hour freedom from most bothersome symptom (MBS) (Table 2).

**Table 1.** Placebo and drug response rates, drug effect, and number needed to treat (NNTs) for 2-hour pain freedom (PF) in women and men.

|  |  |  | Pain Freedom @ 2 h |  |  |  |  |  |  |  |  |  |  |  |  |
| --- | --- | --- | --- | --- | --- | --- | --- | --- | --- | --- | --- | --- | --- | --- | --- |
|  |  |  | Women |  |  |  |  |  |  | Men |  |  |  |  |  |
| Compound | Study | Dose | Placebo* | Drug* | Drug effect (%) | CI (95%) | NNT | P-value |  | Placebo* | Drug* | Drug effect (%) | CI (95%) | NNT | P-value |
| Ubrogepant | 1 | 100 mg, PO | 44/407 (10.8%) | 83/386 (21.5%) | 10.7% | (5.6, 15.8) | 10 | <0.0001 |  | 10/49 (20.4%) | 12/62 (19.4%) | -1.1% | (-16.0, 13.9) | ∞ | 0.8901 |
|  | 1 | 50 mg, PO | 44/407 (10.8%) | 75/379 (19.8%) | 9.0% | (4.0, 14.0) | 12 | 0.0004 |  | 10/49 (20.4%) | 6/43 (14.0%) | -6.5% | (-21.8, 8.9) | ∞ | 0.4151 |
|  | 2 | 50 mg, PO | 58/402 (14.4%) | 93/423 (22.0%) | 7.6% | (2.3, 12.8) | 14 | 0.0050 |  | 7/54 (13.0%) | 8/41 (19.5%) | 6.5% | (-8.5, 21.6) | 16 | 0.3859 |
| Rimegepant | 303 | 75 mg, PO | 62/579 (10.7%) | 128/568 (22.5%) | 11.8% | (7.6, 16.1) | 9 | <0.0001 |  | 12/103 (11.7%) | 14/101 (13.9%) | 2.2% | (-6.9, 11.4) | 46 | 0.6359 |
| Zavegepant | 201 | 10 mg, IN | 54/338 (16.0%) | 78/333 (23.4%) | 7.4% | (1.5, 13.4) | 14 | 0.0153 |  | 8/63 (12.7%) | 10/58 (17.2%) | 4.5% | (-8.2, 17.3) | 22 | 0.4830 |
|  | 301 | 10 mg, IN | 75/546 (13.7%) | 117/506 (23.1%) | 9.4% | (4.7, 14.1) | 11 | <0.0001 |  | 21/100 (21.0%) | 30/117 (25.6%) | 4.6% | (-6.6, 15.9) | 22 | 0.4216 |
| Pooled data (all studies) |  |  | 293/2272 (12.9%) | 574/2595 (22.1%) | 9.5% | (7.4, 11.6) | 11 | <0.0001 |  | 58/369 (15.7%) | 80/422 (19.0%) | 2.8% | (-2.5, 8.2) | 36 | 0.2992 |
| * Number of responders/total number of participants (% of responders); Mantel-Haenszel weighting was used |  |  |  |  |  |  |  |  |  |  |  |  |  |  |  |

**Table 2.** Placebo and drug response rates, drug effect, and number needed to treat (NNTs) for 2-hour most bothersome symptom (MBS) freedom in women and men.

|  |  |  | MBS Freedom @ 2 h |  |  |  |  |  |  |  |  |  |  |  |  |
| --- | --- | --- | --- | --- | --- | --- | --- | --- | --- | --- | --- | --- | --- | --- | --- |
|  |  |  | Women |  |  |  |  |  |  | Men |  |  |  |  |  |
| Compound | Study | Dose | Placebo* | Drug* | Drug effect (%) | CI (95%) | NNT | P-value |  | Placebo* | Drug* | Drug effect (%) | CI (95%) | NNT | P-value |
| Ubrogepant | 1 | 100 mg, PO | 105/405 (25.9%) | 147/386 (38.1%) | 12.2% | (5.7, 18.6) | 9 | 0.0002 |  | 21/49 (42.9%) | 22/62 (35.5%) | -7.4% | (-25.6, 10.9) | ∞ | 0.4285 |
|  | 1 | 50 mg, PO | 105/405 (25.9%) | 145/378 (38.4%) | 12.4% | (5.9, 18.9) | 9 | 0.0002 |  | 21/49 (42.9%) | 17/42 (40.5%) | -2.4% | (-22.7, 17.9) | ∞ | 0.8184 |
|  | 2 | 50 mg, PO | 112/402 (27.9%) | 169/422 (40.0%) | 12.2% | (5.8, 18.6) | 9 | 0.0002 |  | 13/54 (24.1%) | 11/41 (26.8%) | 2.8% | (-15.0, 20.5) | 37 | 0.7595 |
| Rimegepant | 303 | 75 mg, PO | 161/579 (27.8%) | 206/568 (36.3%) | 8.5% | (3.1, 13.8) | 12 | 0.0021 |  | 22/103 (21.4%) | 29/101 (28.7%) | 7.4% | (-4.5, 19.2) | 14 | 0.2252 |
| Zavegepant | 201 | 10 mg, IN | 114/338 (33.7%) | 145/333 (43.5%) | 9.8% | (2.5, 17.1) | 11 | 0.0090 |  | 21/63 (33.3%) | 19/58 (32.8%) | -0.6% | (-17.3, 16.2) | ∞ | 0.9465 |
|  | 301 | 10 mg, IN | 172/546 (31.5%) | 205/506 (40.5%) | 9.0% | (3.2, 14.8) | 12 | 0.0023 |  | 29/100 (29.0%) | 42/117 (35.9%) | 6.9% | (-5.5, 19.3) | 15 | 0.2804 |
| Pooled data (all studies) |  |  | 664/2270 (29.3%) | 1017/2595 (39.2%) | 10.2% | (7.6, 12.9) | 10 | <0.0001 |  | 106/369 (28.7%) | 140/421 (33.3%) | 3.2% | (-3.3, 9.7) | 32 | 0.3349 |
| * Number of responders/total number of participants (% of responders) |  |  |  |  |  |  |  |  |  |  |  |  |  |  |  |

In the female patient population, the three approved gepants for the acute treatment of migraine (ubrogepant, rimegepant, zavegepant) produced a statistically significant drug effect on the 2-hour PF endpoint ranging from 7.4% to 11.8% which corresponded to an average (Mantel-Haenszel weighting) drug effect of 9.5% (CI: 7.4 to 11.6) and an average Numbers Needed to Treat (NNTs) of 11 (9 to 14) (Table 1). Conversely, these three gepants had a markedly lower drug effect on the 2-hour PF endpoint in the male patient population, with drug effects ranging from -6.5% to 6.5% which corresponded to an average drug effect of 2.8% (CI: -2.5 to 8.2) and an average NNTs of 36 (16 to ∞) (Table 1).

Further, in the 2-hour MBS endpoint in the female patient population, the effects of these three gepants ranged from 8.5% to 12.4% which corresponded to an average drug effect of 10.2% (CI: 7.6 to 12.9) and an average NNT of 10 (9 to 12) (Table 2). Conversely, these drugs had a markedly lower drug effect on the 2-hour MBS endpoint in the male patient population, with drug effects ranging from -7.4% to 7.4%, corresponding to an average drug effect of 3.2% (CI: -3.3 to 9.7) and an average NNTs of 32 (14 to ∞) (Table 2).

While these studies were neither designed nor powered to demonstrate efficacy in the male population, the CDER review of the ubrogepant and rimegepant NDAs concluded that “*no therapeutic gain was seen in the male population*”. We therefore reviewed the totality of the result across all approved dose levels. The directionality of point estimates provided in the FDA reviews allow statistical exploration of the results. Under the assumption of no effect between treatment and placebo or no difference between the effects for females and males, there would be a 50% probability that the point estimates would favor either group. Thus, it is possible to compare how often the point estimates favor treatment over placebo or favor females over males to 50%.

In all treatment-dose comparisons for the co-primary endpoints (2h PF and 2h MBS freedom, i.e., 12 total comparisons) the effect was larger in females for every comparison. Under the assumption of no sex difference, a result this extreme would arise in only 1 of 4096 times (p=0.00024); this finding is consistent with a preferential female effect of CGRP-R antagonists in acute treatment of migraine. The CDER data for analysis of the outcomes by sex of gepant effects showed that in each comparison, the female treatment effect was positive, and the resulting p-value is <2% in 100% of cases. In contrast, the p-values for the male treatment effect are >20% in all cases, a finding that is not surprising given the relatively small sample sizes for male subjects. Notably, however, in males nearly half of the treatment effects (2 of 6 for the 2h PF and 3 of 6 for the MBS) favor placebo over active treatment (Table 2 and Fig. 1B, E). While 5 out of 12 estimates in the “wrong” direction does not prove a lack of effect, this would be the expected outcome for a treatment with little to no effect. As noted above, the pooled treatment effect is positive but close to zero for male subjects (2.8% for PF and 3.2% for MBS) (Fig. 1C, F). The similarly pooled treatment effects for female subjects are three times as large at 9.2%, and 10.2%, respectively (Fig. 1C, F).

**Figure 1:**
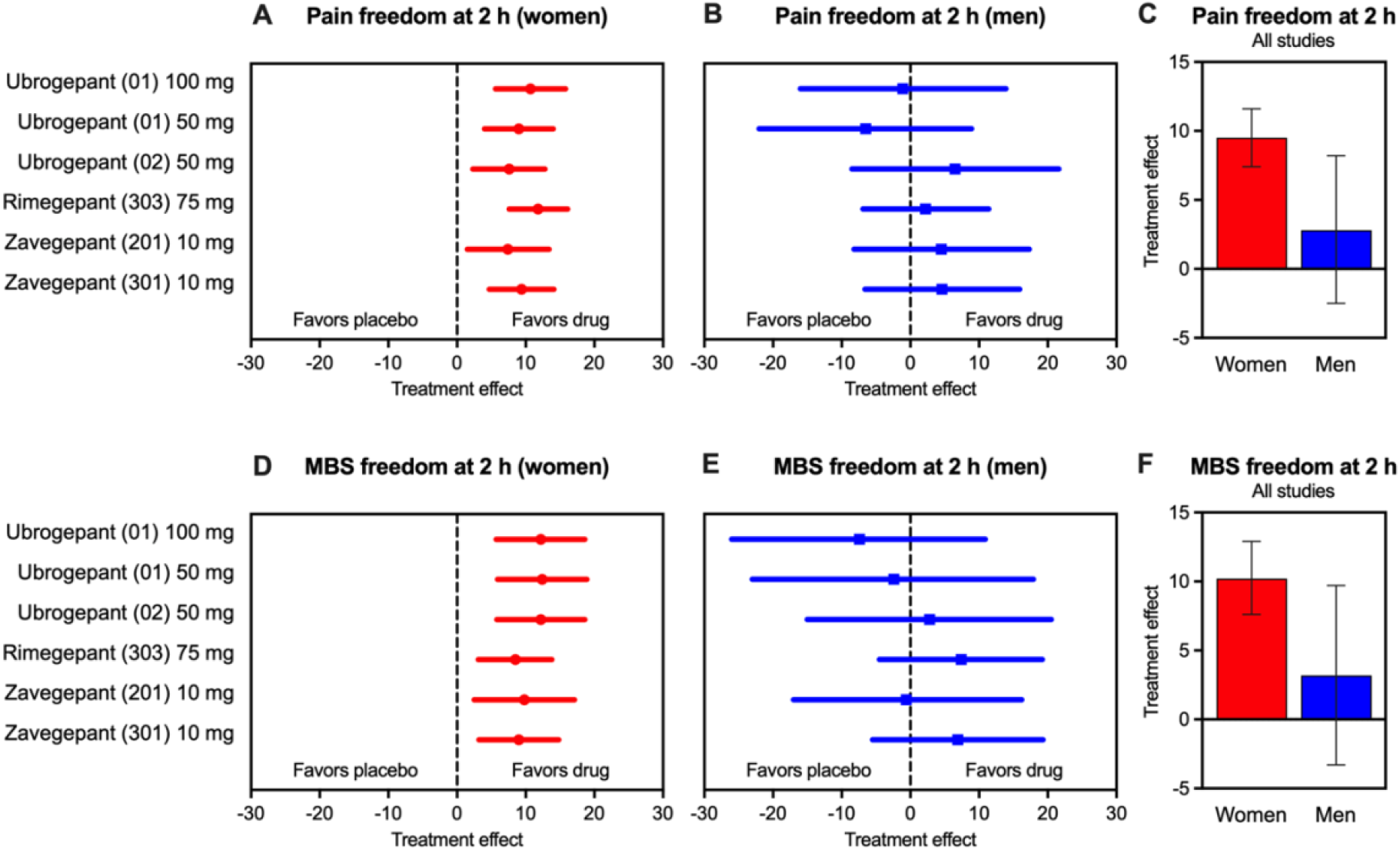
Drug effect of zavegepant, ubrogepant and rimegepant for the acute treatment of migraine in female and male patients. (A) 2-hour PF in six different treatment regiments investigated in the indicated clinical reviews. (B) 2-hour PF combined for all gepant treatments. (C) 2-hour freedom from MBS co-primary endpoint in six different treatment regiments. (D) 2-hour freedom from MBS combined for all gepant treatments. Graphs and analyses were generated using data from the corresponding Center for Drug Evaluation and Research (CDER) clinical reviews for the New Drug Applications (NDA). Data are presented as the mean drug effect size (%) and 95% CI.

### Reasons for larger treatment effects of gepants for acute migraine in women

It is possible that target engagement and pharmacokinetics of these drugs might contribute to the different reported treatment effects in men and women. Dose-response curves for the gepants however, are shallow suggesting that little therapeutic gain is obtained with higher doses and multiple doses of the same drug (i.e., ubrogepant; 50 mg, 100 mg) have been evaluated clinically with no significant improvement of outcomes in men (Table 1). The possibility that women might treat attacks earlier resulting in improved effects also seems unlikely given that improved outcomes are not seen across a range of other therapies including triptans.^11^ It is therefore important to consider the fundamental biological differences between men and women as key factors in the mechanisms that promote migraine as well as in outcomes of migraine therapies.

### Consideration of sex as a biological variable in therapy for acute migraine

The increased female prevalence of migraine begins at the age of menarche, peaks at age 40 and diminishes as women approach menopause.^12,13^ Many women experience increased migraine attacks at or around the time of their menstrual periods, in the first trimester of pregnancy and in the perimenopause. The influence of female hormones to migraine is therefore critical across all life stages experienced by women.^11,14^ The onset of menstrual bleeding co-insides with falling levels of hormones including progesterone and estrogen. Fluctuations in sex hormones have been shown to influence CGRP release and to activate the trigeminovascular system.^15^ Women with regular menstrual cycles have higher CGRP concentrations in plasma and tear fluid.^16^ Women using estrogen containing oral contraceptive pills have CGRP levels similar to women without migraine suggesting that stable estrogen levels may be protective for migraine. Possible differences may thus occur in the efficacy of CGRP-R antagonists in postmenopausal women who generally have lower CGRP levels.^16^

Data from the CDER reviews of gepant drugs are consistent with the likely influence of female hormones across life. Possibly diminished efficacy in promoting PF from acute migraine for ubrogepant and rimegepant was observed in a small number of patients over age 65 (Table 3). While the reported data from these trials were not separated by sex, both trials included a high percentage of women (approximately 85%) suggesting that the decreased effect observed in older patients may represent the contributions of a large group of post-menopausal women (Table 3). Similarly, recent CDER evaluation of zavegepant data reveal diminished effects when studied in patients over the age of 40. In patients younger than 40, zavegepant was significantly more effective in producing 2 hour PF than in those over age 40 (Table 3). This trial also did not separate the data by sex but the population over the age of 40 likely includes a larger proportion of post-menopausal women than those under 40. The smaller effect observed in older patients is therefore consistent with diminished effects of CGRP-R antagonism in postmenopausal women.

**Table 3.** Treatment effect size on the 2-hour PF endpoint and NNT as a function of age.

| Pain Freedom @ 2 hours | Age < 65 years |  |  |  |  | Age ≥ 65 years |  |  |  |
| --- | --- | --- | --- | --- | --- | --- | --- | --- | --- |
|  | Placebo n/N (%) | Drug n/N (%) | Treatment Effect (%) (95% CI)<br>p-value | NNT |  | Placebo n/N (%) | Drug n/N (%) | Treatment Effect (%) (95% CI)<br>p-value | NNT |
| Ubrogepant (50 mg) | 113/889 (12.7%) | 179/860 (20.8%) | 8.1%<br>(4.6%, 11.6%)<br><0.0001 | 13 |  | 6/23 (26.1%) | 3/26 (11.0%) | -14.6%<br>(-36.3%, 7.2%)<br>0.0947 | ∞ |
| Rimegepant (75 mg) | 209/1708 (12.2%) | 347/1714 (20.3%) | 8.0%<br>(5.6%, 10.5%)<br><0.0001 | 13 |  | 6/50 (12.0%) | 4/35 (11.4%) | -0.6%<br>(-14.4%, 13.3%)<br>0.4679 | ∞ |
| Pain Freedom @ 2 hours | Age < 40 years |  |  |  |  | Age ≥ 40 years |  |  |  |
|  | Placebo n/N (%) | Drug n/N (%) | Treatment Effect (%) (95% CI)<br>p-value | NNT |  | Placebo n/N (%) | Drug n/N (%) | Treatment Effect (%) (95% CI)<br>p-value | NNT |
| Zavegepant (10 mg) | 81/522 (15.5%) | 129/485 (26.6%) | 11.1%<br>(6.1%, 16.1%)<br><0.0001 | 9 |  | 77/525 (14.7%) | 106/529 (20.0%) | 5.4%<br>(0.8%, 9.9%)<br>0.0107 | 19 |

### Sexual dimorphism in CGRP-R expression

The female selectivity of CGRP could result from differences in expression or signaling at receptors that bind this peptide. CGRP signals through the canonical CGRP-R, consisting of the calcitonin-like receptor (CLR), a G protein-coupled receptor, and an associated receptor activity modifying protein 1 (RAMP1).^17^ Additionally, CGRP can signal through the amylin 1 (AMY1) receptor that includes RAMP1 along with the G protein-coupled receptor, calcitonin receptor (CTR). Activation of AMY1 which expresses RAMP1 but not CLR, can promote migraine in patients with primary headache disorders.^17^ In contrast, adrenomedullin receptors 1 and 2 (AM1/2) express CLR but not RAMP1 and have not been implicated in inducing migraine. The relative contributions of these receptors to migraine continue to be an area of active investigation. Nevertheless, possible mechanistic insights may be deduced by a comparison of the relative affinities of the gepants for these receptors. Ubrogepant, rimegepant and zavegepant all show comparable low picomolar affinities for the canonical CGRP receptor but vary significantly in their affinities for the AMY1 receptor.^18^ In spite of this, the clinical benefits of gepants are very similar (Tables 1,2 and Fig. 1) suggesting that CGRP signaling promoting migraine likely requires interactions with both CLR and RAMP1. The female selectivity of CGRP might thus be influenced by sexual dimorphism in the expression of either CLR and/or RAMP1. Future studies in preclinical models and in human tissues may help to clarify whether differences in expression of CGRP signaling components can contribute to female selective actions of CGRP and CGRP-R antagonists in migraine.

### Prolactin, CGRP and migraine

The actions of CGRP in migraine are influenced by multiple sex hormones. Recent studies highlight the role of prolactin, a circulating neurohormone that is released by the pituitary and produced locally by multiple cell types. Significantly, this female predominant hormone enhances CGRP release and has a demonstrated role in migraine.^19^ Prolactin is under control of estrogen as well as stress. Women (and female animals) have higher circulating levels of prolactin than males as well as greater stress responses. Prolactin is released from pituitary lactotrophs and is under tight inhibitory control by hypothalamic dopaminergic tuberoinfundibular (TIDA) cells. Preclinical studies show that the TIDA cells are inhibited by stress-related transmitters resulting in increased circulating prolactin.^20^ Dopamine D2 receptor agonists are used therapeutically to reduce release of prolactin from the pituitary. In humans, stress increases circulating prolactin, lowers sensory thresholds, is associated with painful menstruation (i.e., dysmenorrhea) and importantly, is the most common self-identified migraine trigger.^21^ Patients with very high levels of prolactin resulting from pituitary adenomas have increased migraine attacks that are resolved with treatment by dopaminergic agonists that reduce prolactin levels.^22^ Estrogen and stress-related influences on prolactin are therefore likely to have a CGRP component in promoting migraine attacks in women.

### CGRP and prolactin are profoundly female-selective in their actions

Preclinical studies show that neurotransmitters can be sexually dimorphic in their effects. In preclinical models, CGRP produces female-selective pain and headache responses.^23^ While rodent models cannot capture the complex multi-symptom and multi-phasic nature of migraine, they can and do provide mechanistic information that helps to explain migraine symptoms. As migraine headache likely arises from trigeminal nociceptors that innervate the cranial meninges, direct activation of these afferents in animals provides a surrogate measure of headache through assessment of pain behaviors including cutaneous cephalic allodynia, also observed in patients during migraine attacks. Dural stimulants also elicit other surrogate measures including facial grimace in rodents that may be representative of ongoing pain. Importantly, application of CGRP directly to the dura mater elicited both evoked and ongoing migraine-like pain behaviors at much lower doses and with longer lasting effects in female, compared to male mice.^23^

Preclinical studies have consistently demonstrated that prolactin produces selective sensitization of female nociceptors that is associated with increased release of CGRP.^24^ Application of prolactin to the mouse dura mater produces headache-like pain behaviors in female, but not male, mice that is blocked by either a prolactin receptor antagonist or by a CGRP receptor antagonist.^19^ These studies reveal cross-talk between prolactin and CGRP that is relevant to migraine like pain in females.^19^ Stress-induced sensitization of trigeminal ganglion (TG) afferents to a TRPA1 agonist that has been linked to headache in humans has also been shown to be blocked by dopaminergic agonists or by interference with prolactin signaling at prolactin receptors selectively in female mice.^20^

### Sexual dimorphisms in rodent and human nociceptors

Recent discoveries have challenged our long-held understanding of somatosensory mechanisms promoting the perception of pain and headache. Nociceptors are sensory fibers with cell bodies residing in dorsal root ganglia (DRG) and trigeminal ganglia that detect and transmit high intensity stimuli capable of eliciting actual or potential tissue damage from the peripheral tissues to the central nervous system. Remarkably, analyses of human post-mortem DRG neurons revealed that these cells are sexually dimorphic.^25^ Thus, nociceptors, the fundamental building blocks of pain are different in men and women and, by extension, the mechanisms that may promote pain can also differ between the sexes.

A key observation is the sexual dimorphism in transcript for CGRP that is expressed at higher levels in DRG neurons recovered from human female, compared to male, donors.^25^ Additionally, transcriptome analysis of non-neuronal satellite cells in post-mortem human DRG have also been found to differ between male and female tissues suggesting additional sexually dimorphic mechanisms that may promote neuronal activation and signaling.^26^ Most recently, sexual dimorphism has been observed in human nociceptors at the protein and functional level.^27^ Prolactin receptors are expressed at higher levels in DRG cells recovered from women. Furthermore, incubation of nociceptors from female, but not male, human donors with prolactin produced increased excitability of the neurons. These studies support the conclusion that there are male and female nociceptors that can be distinguished at the transcript, protein and functional levels. While limited by assessment of a very small sample size, it appears that similar conclusions can be drawn from analysis of human TG neurons.^28^ Corresponding findings from the analysis of human tissues and from preclinical studies support sexual dimorphism in nociceptors and the profound female selectivity of both CGRP and prolactin. Such information suggests an opportunity for the discovery and implementation of therapies for treatment of pain and migraine based on patient sex.

## Conclusions

Clinical data support the conclusion that CGRP-based therapies are female selective and effective in women for acute migraine therapy. The relative absence of data that demonstrate clinical benefit of gepant therapies for acute migraine treatment in men suggests the need for a careful consideration of decision making in the choice of care and for discussion and patient disclosure. Additionally, to establish efficacy in men, dedicated and appropriately powered clinical trials should be performed. Similarly, efficacy estimates should be established for pre- and postmenopausal women separately.

Collectively, new information from preclinical studies and clinical data supports the conclusion that while the criteria for fulfillment of a migraine diagnosis may be met by men and women, there is a likely qualitative sex difference where underlying pathophysiological mechanisms including the contribution of CGRP to migraine may differ between the sexes. These findings with migraine also raise the possibility that conclusions as to the efficacy of CGRP-based therapies for other, non-migraine, but female prevalent pain conditions such as fibromyalgia, irritable bowel syndrome and others could have been influenced by the relative proportion of men and women, as well as the percentage of pre-vs. postmenopausal women, in any given trial. Overall, our conceptual view of therapy for migraine and other pain conditions requires adjustment so that studies consider patient sex as the most fundamental aspect of precision medicine.

## Data Availability

All data produced in the present study are available upon reasonable request to the authors

